# Risk-taking behavior and sustained attention after a night shift: an observational study in healthcare professionals

**DOI:** 10.1101/2021.02.15.21251795

**Authors:** S. Bruno, F. Cruz-Sanabria, A. Bazzani, P. Frumento, U. Faraguna

**Author notes:** corresponding author: U. Faraguna, mail; via San Zeno, 31, 56123, Pisa, Italy. Department of Translational Research and of New Surgical and Medical Technologies – University of Pisa - Pisa, Italy; Department of Developmental Neuroscience – IRCCS Fondazione Stella Maris – Pisa, Italy.

## Abstract

**Study Objectives:** Night work plays an irreplaceable role in healthcare systems. However, sleep loss due to night shifts can negatively affect healthcare workers’ cognition. Identifying individual variables predicting night shift tolerance may help to mitigate the negative consequences of sleep loss on workers’ and patients’ safety. This study aims to explore, in a healthcare workers’ sample, which variables predict the possible impairment in sustained attention and increase in risk-taking behavior after a night shift.

**Methods:** 22 healthcare workers (avg. age 39.4 ± 11.5 y, 63% females) participated in the experiment. Participants wore a wrist actigraph during a night shift and the preceding 6 days. At the beginning (t0) and at the end (t1) of the night shift, they underwent an assessment of sustained attention (Psychomotor Vigilance Task, PVT) and risk-taking behavior (Balloon Analogue Risk Task, BART). Questionnaires on risk propensity (Domain-Specific Risk Taking, DOSPERT) and on demographics were also administered. Linear regression models were estimated to disentangle the effect of age, risk propensity and actigraphically-defined sleep metrics on cognitive variables.

**Results:** Wake after sleep onset (WASO) significantly predicted t1-t0 PVT metric “major lapses” (β=-0.11, p=0.0007) while age and DOSPERT significantly predicted t1-t0 BART score (β=-0.26, p=0.023 and β=-0.097, p=0.043, respectively).

**Conclusions:** Sleep parameters predict the impairment in sustained attention, while the combination of age and risk propensity predicts an increase in risk-taking behavior. The implementation of measures to prevent cognitive decline during night shifts should be designed according to the type of tasks workers must accomplish.

## BACKGROUND

Shift work is common in many professions. Worldwide, approximately 20% of workers experience night shifts outside the daytime 07:00 h to 18:00 h window.[1] Night work is thus a major concern for a significant share of working industries: indeed, it has been associated with both reduced working and cognitive performance[2,3] and increased risk of developing several medical conditions, among which sleep disorders are the most frequent.[4] Moreover, the risk of occupational injury is increased during night work compared to conventional day shifts.[5,6]

From a chronobiological perspective, night work forces human beings to stay awake during an unfavorable time of day, causing circadian misalignment and cumulative sleep deprivation.[7,8] Sleep loss is accountable for both reduced working performance[9,10] and the health issues shift workers are more likely to experience.[11] Moreover, it is well known that the dysregulation of sleep/wake pattern leads to the impairment of multiple executive functions,[12] thus contributing to the decline in cognitive performance during the night shift.[13] In healthcare systems, impaired cognitive functioning of night workers can contribute to the explanation of the increased occurrence of medical errors during off-hours[14–16] resulting in a threat to patient safety.[17] Predicting individual night work tolerance might help mitigate night shifts’ negative impact on both workers and patients.

Among executive functions, sustained attention is probably the most extensively studied in relation to sleep loss.[18] Sustained attention is defined as “a state of readiness to detect rarely and unpredictably occurring signals over prolonged periods of time”[19] Sleep deprived individuals show slower response speed and higher numbers of commission and omission errors than well rested subjects.[20] Moreover, the impairment in more complex cognitive tasks, reported under sleep deprivation condition, may be a consequence of participants’ inability to focus their attention on the task at hand for a sufficient time frame, although several studies suggest that sleep loss impacts higher cognitive functions irrespective of vigilance.[18] The impact of night work on sustained attention was well investigated in healthcare workers (HCWs). Night shift nurses show higher reaction times (RTs) compared to daytime shift nurses. Moreover, the mean of the fastest 10% RTs is higher at the end of a night shift than the pre-shift baselines.[21] HCWs show higher mean RT (i.e. they are slower) during night shift compared to daily shifts,[22] while attentional performance gradually deteriorates throughout working nights.[23] However, inconsistently with the aforementioned studies, Reinke and colleagues[24] and Capanna and co-workers[25] observed no effect of night work on sustained attention. Individual variability in night shift work tolerance may explain these conflicting results. The definition of shift work tolerance (SWT) is the ability to adapt to shift work without adverse consequences.[26] Many variables have been addressed to contribute to different SWT in terms of work-related sleep disturbances and cognitive performance.[27–29] Among those, older age is associated with poor night shift attentional performance,[13] whereas a sufficient sleep duration during the night shift can grant a partial recovery of attentional impairment.[30]

Along with sustained attention, risk-taking too is influenced by sleep deprivation.[12,31] Risk-taking as a cognitive construct may be explored through two different approaches: as a personality trait or as the likelihood of contingently engaging in risky behavior.[32] Individual differences in propensity to risky choices, as a stable personality trait, are addressed as differences in risk attitude and have been investigated trough validated questionnaires such as the Domain-Specific Risk-Taking questionnaire (DOSPERT).[33] Risk-taking behavior (RTB), instead, is defined as “an action or decision that has both the potential for (a) danger, loss, or harm, and (b) the gain of some form of reward”.[34,35] Unlike risk attitude, RTB is not stable across situations:[36] e.g., individuals show different degree of risk propensity in financial, recreational or other domains.[33] Moreover, both under controlled laboratory condition and during field experiments, RTB has proven sensitive to sleep loss,[34] even if only a few studies focused on night shifts’ impact on RTB in the healthcare context. Those who did it showed, in two different samples of relatively young residents, an increase in RTB after a 24-hour call,[37] as well as after three consecutive working nights in an emergency unit.[25] To the best of our knowledge, no study tried to replicate these findings in elder professionals. The available results about the effects of night shifts on RTB obtained in young professionals cannot be automatically extended to older colleagues, as ageing tends to decrease both risk attitude[38] and RTB.[39] In parallel, studies involving forced desynchrony and sleep restriction protocols showed that cognitive performance is more stable across different experimental conditions in elder as compared to younger participants.[40–44] Such results suggest that the ability to cope with sleep/wake disruptions increases with ageing, thus making it difficult to disentangle the effect of age, sleep deprivation and risk attitude on the night work-related RTB increase.

In healthcare workers (HCW) populations, attitude toward risk explains some of the variability in clinical practice.[45] Allison et al.[46] demonstrated that higher risk propensity in medical decision-making is associated with lower patients’ charge, while a study from Pearson and colleagues[47] showed that emergency care patients’ rate of admission was lower when professionals with high scores in a risk-taking scale were on shift. High risk attitude has also been associated with low adherence to safety procedures, thus increasing the risk of workplace accidents.[48] More recently, risk tolerance has been addressed as a predictor of medical decision-making.[49] Risk attitude-exploring questionnaires have never been used as profiling tools to identify professionals whose decision-making is more likely to turn riskier after a night shift. Indeed, individual variables associated with a reduced impact of night working on RTB have not been so far identified. Consequently, it cannot be argued that variables associated with an improved shift work tolerance in terms of sustained attention also confer an improved shift work tolerance in terms of RTB. The aim of the study is twofold:

1. to explore whether night shift’s impact on risk-taking behavior and sustained attention in healthcare workers is predicted by age, risk attitude and night shift sleep-related actigraphic parameters;
2. to explore whether age, risk attitude or night shift sleep-related actigraphic parameters specifically predict night shift impact on RTB and sustained attention.

## METHODS

### Study design and procedure

Six days before the night shift during which the experiment took place, participants were provided with a watch-shaped wrist actigraph bracelet. They had to wear it on the non-dominant arm for seven days, following their daily routines until the end of the night shift. Participants also underwent two computer-based cognitive tests at the beginning (t0) and at the end (t1) of the night shift: the Psychomotor Vigilance Task (PVT) and the Balloon Analogue Risk Task (BART). The two tasks were performed in the same order by all participants both at t0 and t1. The cognitive assessments were performed in a quiet room within the hospital ward at the same time of the day (t0: 6-8 p.m.; t1: 7-9 a.m.), in order to minimize the variability of circadian oscillations on participants’ cognitive performance.[50] Participants also completed a socio-demographic questionnaire and the Domain Specific Risk-Taking scale (DOSPERT). The study has received the approval of the Bioethical Committee of the University of Pisa on August the 30th, with protocol number 0098770/2019. All participants provided written informed consent before joining the study. Data collection started in September 2019 and stopped in January 2020.

### Participants

Twenty-four healthcare practitioners (twenty-two physicians and two laboratory technicians; avg. age 39.4 ± 11.5 y, 63% females) working in ten different wards agreed to participate in the study. Their working schedules included daily 6-h shifts (8 a.m – 2 p.m.; 2 p.m. – 8 p.m) and 12-h night shifts (8 p.m. – 8 a.m.). Inclusion criteria were full-time working and performing at least one night shift per month. Research exclusion criteria were: suffering for neurological, psychiatric or serious medical conditions, as well as from sleep disorders; the use of drugs that may interfere with sleep or decision-making process; having travelled in another time zone in the two weeks preceding the experiment’s night shift. One participant was ruled out from the study because of a malfunctioning of the actigraphic device (sleep/wake data were not available) and another was removed from the sample because of the low quality of the cognitive assessments. Night shift’s sleep data of two other subjects were missing due to unexpected, anticipated end of actigraphic recording: analyses on sleep data were thus performed on 20 observations.

### Materials

#### Sociodemographic and work characteristics

Demographics and work-related variables included: age; gender; Body-Mass Index (BMI); alcohol, cigarettes and coffee consumption; hospital department; working position; avg. weekly working hours; avg. monthly night shifts.

#### Actigraphic measures

Participants continuously wore a waterproof wrist actigraph Fitbit Flex2 (FF2) on their non-dominant wrist for 7 days, with no interruption. Data were sampled in 1-minute epochs and digitally stored for subsequent analysis. The following quantitative sleep parameters derived from the FF2 were estimated through a validated and certified artificial neural network (ANN)-based algorithm:[51,52]

- Total Sleep Time (TST), defined as the total duration of sleep (in hours and minutes);
- Waking After Sleep Onset (WASO), defined as the time spent awake after sleep onset and before sleep offset (in minutes);
- Sleep Efficiency (SE), defined as the time spent asleep between sleep onset and offset (in %).

#### Cognitive tasks

The Balloon Analogue Risk Task (BART) is a computer-based behavioural measure of RTB.[53] Participants are asked to inflate simulated balloons which are shown on the computer screen. By pushing the pump button, participant accrue 5 cents temporarily. If the balloon is excessively pumped, it explodes, and the money stored in the temporary reserve is lost. Three types of balloons differing in their explosion point are randomly shown: type 1 yellow balloons, with an average life of 4 pumps; type 2 orange balloons, with an average life of 16 pumps; type 3 blue balloons, with an average life of 64 pumps. No hint is provided to participants about the color code. At any point during each trial, participants can click the “Collect $$$” button to transfer all the money from the temporary reserve to a permanent one. After each balloon explosion or money collection, a new balloon appears on the screen and a new trial begins. The test ends after 90 trials. BART scores were computed both as mean number of pumps per balloon and adjusted number of pumps per unburst type 3 balloon.

The Psychomotor Vigilance Task (PVT) is a reaction time test.[54] Participants sit comfortably in front of the computer and are asked to respond to random visual stimuli (red circles) which appear in the center of a dark computer screen with an inter-stimulus interval (ISI) varying between 1 and 9 seconds. They are also told to avoid pressing the spacebar when no stimulus is shown on the screen: if this happens, it is counted as a false start (or anticipation or commission error). Response times greater than 500 ms are called “major lapses” (or omission errors) and are considered a hallmark of sleep deprivation.[20] The whole experiment lasts 10 minutes.

In order to describe the impact of a night shift on HCW cognitive performance, differential indices were computed by subtracting the behavioral tasks scores performed by each participant at t0 from the score performed at t1. These metrics are hereby labelled as t1-t0 scores.

The cognitive tasks were run using the Psychology Experiment Building Language (PEBL), a free open-source software that allows researchers to design and run cognitive tests.[55]

#### Domain Specific Risk-Taking questionnaire (DOSPERT)

The 30-item questionnaire assesses risk propensity across five different domains: financial decision (separately for investing versus gambling), health/safety, recreational, ethical and social decisions.[33] In the first part, participants rate how probable is their engagement in risky activities using a number between 1 and 7; in the second optional part, respondents are asked to rate the magnitude of the risk and the expected benefits for the same activity judged in part I. The first part global score (probability to be involved in risky activities) was used in the current study as the measure of individual risk attitude. Italian translation published by the authors in the official website of the scale was used in the present study (https://sites.google.com/a/decisionsciences.columbia.edu/dospert/translating-the-scale).

### Statistical analyses

R 3.3.3 GUI 1.69 was used for data analysis. Descriptive statistics were reported with means, standard deviations, medians and boundaries of interquartile range values for continuous variables, percentages for categorical variables, if not specified otherwise. Linear regression models were estimated to assess the effect of age and gender on baseline (t0) sustained attention and RTB and to assess the effect of age, risk propensity and night shift sleep-related actigraphic parameters on t1 sustained attention and RTB scores. The selected sample size (n=22) was sufficient to achieve a power of 0.8 in a paired t-test, assuming that the standardized difference between the means of the two groups was at least 0.6, with a two-sided level of significance of 0.05. All statistical tests were two-sided, and the level of significance was set at 0.05.

## RESULTS

22 HCWs were enrolled in and concluded the study. The corresponding descriptive statistics of demographics, lifestyle, work-related and sleep data are summarized in Table 1.

**Table 1.**
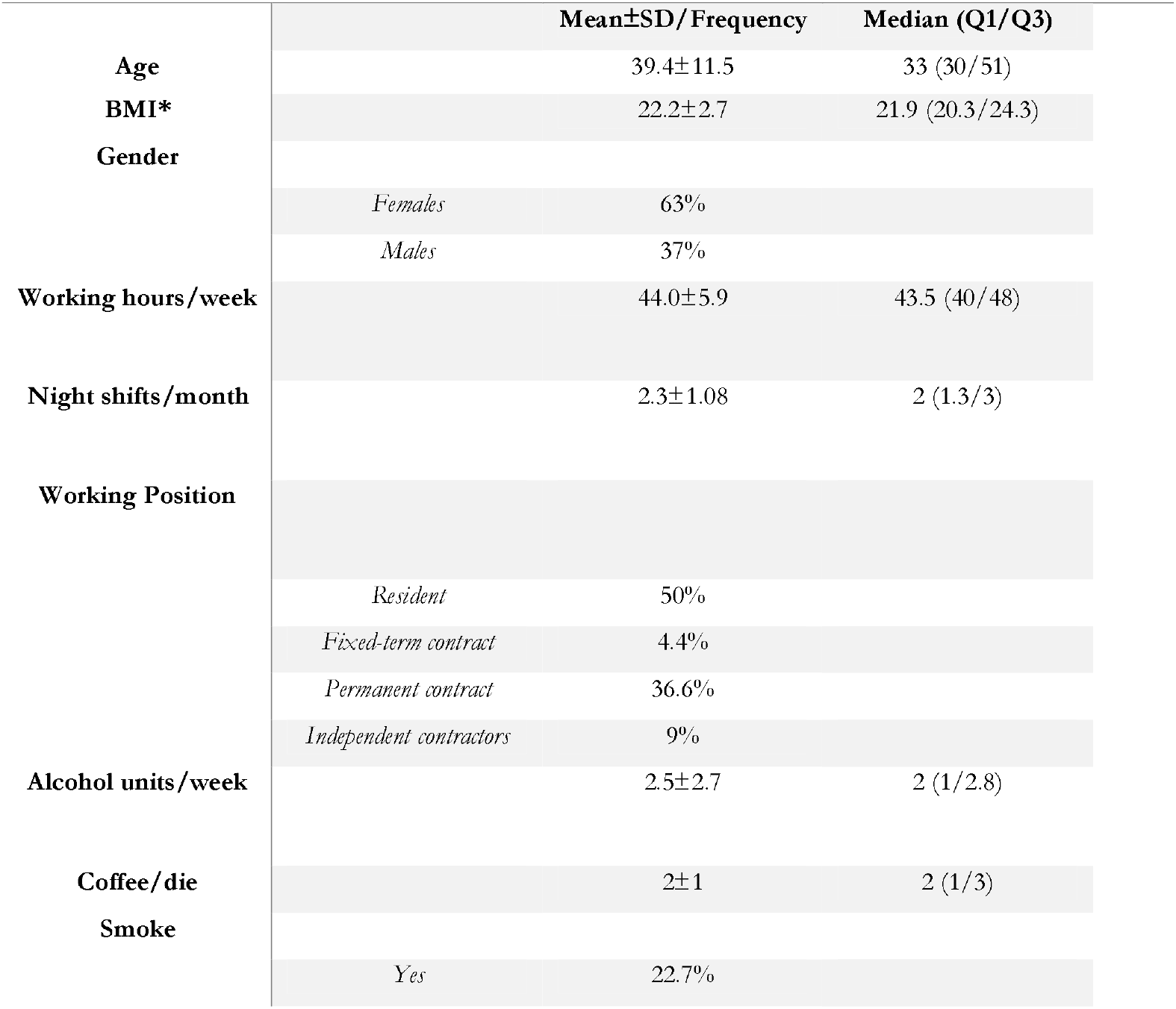
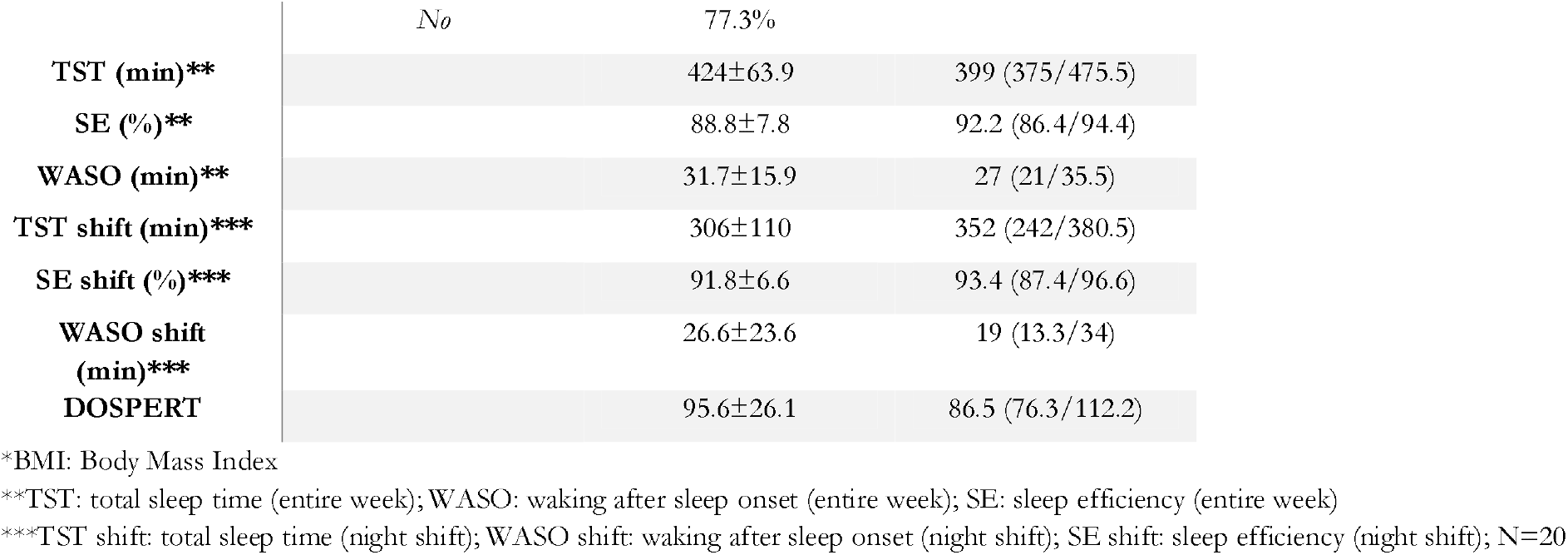
Descriptive statistics of demographics, lifestyle, work-related and sleep data (N=22)

During the experimental night shift, actigraphically-defined mean TST was 306 ± 110 minutes and it significantly differed from the mean TST of the entire week (424 ± 63.9 minutes vs 306 ± 110 minutes, Wilcoxon test, p < 0.001).

Mean values of response time, anticipation and major lapses for PVT and average raw and adjusted number of pumps per balloon for BART were computed at t0, t1 and for t1-t0 (Table 2).

**Table 2.** Descriptive statistics of cognitive data (N=22)

|  | t0 |  | t1 |  | t1-t0 |  |
| --- | --- | --- | --- | --- | --- | --- |
|  | Mean±SD | Median (Q1/Q3) | Mean±SD | Median (Q1/Q3) | Mean±SD | Median (Q1/Q3) |
| Mean response time* (ms) | 388±69.5 | 377 (346.8/398.3) | 389±57.3 | 391 (353/417.3) | 1.8±59.9 | 10 (-10.5/17.6) |
| Anticipations* | 1.4±2.2 | 1 (0/1.8) | 1.1±1.82 | 0.5 (0/1) | -0.3±1.8 | 0 (-1/0.8) |
| Major lapses* | 5.2±7.6 | 3 (2/4.8) | 6.6±6.6 | 4 (2.3/8) | 1.5±3.8 | 1 (0/2.8) |
| Number of pumps** | 10.3±5.7 | 8.5 (6.3/14) | 11.6±5.5 | 12 (7.3/16.3) | 1.3±4.5 | 0.5 (-1/2.8) |
| Adjusted number of pumps** | 17.7±12.5 | 13 (10/25.8) | 22.4±13.2 | 25 (11/30.5) | 4.7±11.1 | 2.5 (-1.8/6) |

\*: PVT scores
\*\*: BART scores

To exclude age- and gender-related baseline differences in sustained attention and RTB, linear regression models were estimated considering t0 PVT metrics (mean response time, anticipations, major lapses) and BART metrics (mean number of pumps per balloon and adjusted mean number of pumps per unburst type 3 balloon) as dependent variables, age and gender as independent variables. Older age significantly predicted a higher number of major lapses (β=0.31 [95% CI: 0.030, 0.59], p=0.030). All other predictors did not reach significance (Results not shown).

With the aim of exploring the impact of night shift sleep-related actigraphic parameters on sustained attention and RTB, five linear regression models were fitted considering as dependent variables t1-t0 PVT and BART metrics; and as independent variables TST, WASO and SE during the night shift. WASO significantly predicted the difference in the number of major lapses between t1 and t0 (t1-t0; β=-0.098 [95% CI: -0.18, -0.011], p=0.029). No other independent variable reached significance (Results not shown).

To disentangle the effect of age, risk attitude and night shift sleep-related actigraphic parameters on sustained attention and RTB, three linear regression models were estimated considering t1-t0 PVT number of major lapses and BART metrics as dependent variables; age, DOSPERT score and WASO as independent variables. t1-t0 number of major lapses was significantly predicted by WASO (β=-0.11 [95% CI: -0.16, -0.052], p=0.0007) but neither by age (β=0.061 [95% CI: -0.075, 0.20], p=0.36) nor by risk attitude (β=-0.009 [95% CI: -0.067, 0.049], p=0.75) (Table 3).

**Table 3.** Linear regression analysis assessing the effect of age, risk attitude and WASO during the night shift on t1-t0 major lapses

|  | Estimate<br>(95% Confidence interval) | Std. Error | Pr(> \|t\|) |
| --- | --- | --- | --- |
| **(Intercept)** | 2.80<br>(-6.44/ 12.04) | 4.36 | 0.53 |
| **Age** | 0.061<br>(-0.075/0.20) | 0.064 | 0.36 |
| **DOSPERT** | -0.009<br>(-0.067/0.049) | 0.027 | 0.75 |
| **WASO (min)** | -0.11<br>(-0.16/-0.052) | 0.025 | 0.0007*** |

WASO: night shift waking after sleep onset
Significance codes: \* $<0.05$ , \*\* $<0.01$ \*\*\* $<0.001$
Dependent variable: t1-t0 major lapses.
N=22 (WASO N=20)
Multiple $R^2=0.53$ ; Adjusted $R^2=0.45$

t1-t0 mean number of pumps per balloon was significantly predicted by age (β=-0.26 [95% CI: - 0.48, -0.041], p=0.023) and risk attitude (β=-0.097 [95% CI: -0.19, -0.004], p=0.043), but not by WASO (β =-0.026 [95% CI: -0.11, 0.060], p=0.53) (Table 4).

**Table 4.** Linear regression analysis assessing the effect of age, risk attitude and night shift quanti-qualitative sleep index on t1-t0 mean number of pumps per balloon

|  | Estimate<br>(95% Confidence interval) | Std. Error | Pr(> \|t\|) |
| --- | --- | --- | --- |
| **(Intercept)** | 21.55<br>(6.63/ 36.47) | 7.038 | 0.0075** |
| **Age** | -0.26<br>(-0.48/-0.041) | 0.10 | 0.023* |
| **DOSPERT** | -0.097<br>(-0.19/-0.004) | 0.044 | 0.043* |
| **WASO (min)** | -0.026<br>(-0.11/0.060) | 0.041 | 0.53 |

WASO: night shift waking after sleep onset
Significance codes: \* $<0.05$ , \*\* $<0.01$
Dependent variable: t1-t0 mean pumps per balloon.
N=22 (WASO N=20)
Multiple $R^2=.37$ ; Adjusted $R^2=.26$

t1-t0 adjusted number of pumps was significantly predicted by age (β=-0.58 [95% CI: -1.07, - 0.085], p=0.024) and nearly significantly predicted by risk attitude (β=-0.20 [95% CI: -0.42, 0.014], p=0.065), but only after the removal of WASO as covariate (Results not shown).

Despite the few observations, residuals followed a bell-shaped distribution. No influential points or outlier were found.

## DISCUSSION

With the present study we explored whether the impact of night shift on risk-taking behavior and sustained attention in HCWs can be explained by specific parameters, i.e. night shift sleep quantity and quality, age, and individual propensity to risk as measured by the DOSPERT. A higher WASO value significantly predicted participants’ sustained attentional impairment but not risk-taking behavior increase, whereas age and risk attitude significantly predicted increasing in risk-taking behavior but not attentional impairment. These results suggest that the impact of a night shift on different cognitive domains may be modulated by different individual variables.

Sleep is essential to ensure a global proper brain functioning: RTB and sustained attention are two of the cognitive domains affected by dysregulation in sleep/wake pattern.[56,57] Sleep deprivation impairs different brain systems through different pathways.[12,58] Focusing on RTB and sustained attention, several studies reported that sleep loss may selectively impact risky decision-making while vigilance is preserved.[25,59] Moreover, the use of stimulants, e.g. caffeine, restores sustained attention in sleep-deprived subjects without modifying their risk assessment ability.[60] Individual vulnerability to the impact of sleep deprivation on cognition should be hence analytically addressed to identify which variables better predict the worst consequences on functions like sustained attention and RTB. This becomes particularly relevant in clinical decisions. Profiling HCWs who frequently face night shifts might help to early detect those workers that are more vulnerable to the negative effects of sleep deprivation on their real-life tasks, so that policy makers and professionals could hopefully counteract the decline in healthcare service quality during night-time.[17]

Our first hypothesis stated that sleep alterations due to night shifts would increase risk-taking behavior and decrease sustained attention in healthcare workers. Also, we hypothesized that age, risk propensity and sleep parameters recorded during the night shift could predict an increase in RTB and a decrease in sustained attention after a night of work. WASO predicted participants’ attentional impairment following a night of work, as measured by t1-t0 number of omission errors while performing PVT. The association between WASO and sustained attention was such that the higher was the WASO value, the lower was the attention impairment after the night shift. These results suggest that a better attentional performance is achieved when sleep is more fragmented. To explain this counter-intuitive result, we hypothesized that, being WASO defined as the number of minutes spent awake after sleep onset and before sleep offset, the metric would not be fully independent of sleep duration: indeed, a longer sleep duration might increase the probability of experiencing brief waking episodes between sleep onset and offset. In order to confirm this hypothesis, a linear regression model was run considering WASO measured during the night shift as dependent variable and TST measured during the night shift as independent variable. As expected, TST significantly predicted WASO (β=0.10 [95% CI: 0.008, 0.20], p=0.034) so that an increased TST was associated with an increased WASO (Results not shown). Another possible explanation is that a higher fragmentation of sleep, reflected by a higher WASO value, led to an increased sleep inertia during t1 assessment,[61] which in turn was reflected by an attentional impairment.[62]

Being significantly associated with WASO, the number of major lapses was confirmed as a PVT metric sensitive to sleep impairments.[20] Previous studies reported an increase also in mean response time after a night shift.[22] In the current study, WASO did not significantly predict t1-t0 mean response time instead. However, the findings are in line with previous works that showed that sleep deprivation does not completely compromise human vigilance ability: rather, sleep loss makes it difficult to maintain a stable attentional performance over time. Sleep-deprived subjects show a high variability in PVT performance alternating major lapses and normal timely responses. According to the “state instability” hypothesis, fluctuations in sustained attention under forced waking conditions are the result of a trade-off between sleep initiating mechanisms and subjects’ effort to keep staying awake.[63] Therefore, the number of major lapses is probably the PVT metric most sensitive to sleep deprivation, given its ability to detect the intermittent attentional deficits that characterize sleep-deprived humans’ cognitive performance. It is thus plausible that, when sleep deprivation is partial and not total, like during HCWs’ night calls, sleep impairments might be accountable for an increase in the number of major lapses despite no change in mean response time.

Unlike sustained attention, no relationship linked WASO to t1-t0 BART scores. Instead, t1-t0 mean number of pumps per balloon was significantly predicted by age and risk attitude. t1-t0 adjusted mean number of pumps per unburst type 3 balloon, instead, was significantly predicted by age and nearly significantly predicted by DOSPERT score only if WASO was not used as covariate. Small sample size may be accountable for the inconsistency between predictions of the two BART metrics. Younger and less risky HCW were therefore more likely to pay the consequences of night working in terms of increased RTB. The result is in line with previous studies outlining RTB increases in residents’ samples following night shifts.[25,37] This result might be interpreted as an increased vulnerability to sleep/wake disruption showed by young humans, consistently with previous in lab studies.[40,41] However, field experiments associated older age with poorer shift-work tolerance,[28] even if some authors claim that limited evidence supports a reduced shift work tolerance in elder workers.[64] Another explanation is that the decreased chance of engaging in a risky behavior associated with ageing[38,39] prevents elder HCW from increasing their RTB under triggering conditions, such as work-related sleep deprivation. Furthermore, individual risk attitude was negatively correlated with the RTB post-shift increase. Two different explanations could help interpreting these results. First, a ceiling effect might preclude risk-seekers from further increasing their RTB under conditions that raise the probability to engage in risky behaviors, e.g. sleep deprivation. Second, the decision of reducing the time dedicated to sleep can be considered a risky behavior by itself, since it is likely to compromise daytime functioning. According to this hypothesis, risk-seekers would better tolerate sleep loss compared to risk-avoiders, because they would be more likely to experience it during their routine life. As a result, the decision-making of risk-avoiders only becomes riskier when elicited by night working, given the unusual condition of sleep deprivation they are exposed to. The association between higher risk propensity and sleep/wake pattern disruption reported by O’brien and Mindell[65] may support the assumption according to which risk-takers are more used to sleep deprivation and therefore able to tolerate it better compared to risk-avoiders. Moreover, a study from Rusnac et al.[66] demonstrated that chronic sleep deprivation is associated with increased RTB and higher scores on a sensation-seeking scale only in subjects who voluntary reduce sleep duration compared to subjects who reduce it involuntarily, i.e. suffering from insomnia. Indeed, no definitive conclusion can be drawn grounded on the current findings: further investigations will shed light on the relationship that links sleep deprivation, risk attitude and risk-taking behavior.

Taken together these findings support the hypothesis according to which the paths leading from night shift to increased RTB and worse sustained attention diverge,[12] thus supporting also our second hypothesis. Indeed, we expected that the variables identified as predictors of the impact of a night shift on human cognition might be valid for one variable, e.g. RTB, but not for the other, i.e. sustained attention. This might also provide an explanation for the heterogeneity and inconsistencies emerged from previous studies on night work. Some authors reported a negative impact of night shift on both sustained attention and RTB,[37] while some others demonstrated instead an increase in RTB not paralleled by an impairment in sustained attention.[25] In this second case, a sufficient sleep quantity and quality during the night shift might have masked the impact of night work on vigilance.

If further studies would confirm the present findings, public health policy makers might consider the implementation of specific strategies to limit the negative effects of night working on clinical decision-making and cognitive processes affected by sleep loss. Indeed, well-established and immediately applicable strategies like napping or the use of caffeine would restore, at least partially, workers’ ability to focus on a repetitive task for a certain time window (i.e. sustained attention performance), but they might be insufficient to prevent vulnerable HCWs risky decisions (i.e. RTB). At an organizational level the quality and safety of healthcare services could benefit from rescheduling tasks and shifts grounded on individual psychometric profiling methods.

To the best of our knowledge, this is the first study aimed at profiling HCWs according to their risk attitude as a trait and exploring contingent RTB vulnerability to night shift in a sample comprising not only young residents. Another strength point is the two-cognitive assessment protocol, which allowed to create a differential t1-t0 index useful in reducing the interindividual variability in the evaluation of cognitive performance. Moreover, actigraphy provided objective measures of sleep in a minimally invasive way, which can be easily transferred to real-life settings to ensure a continuous monitoring of HCWs performance and safety.

The major limitation of this study is the small sample size. The numerosity of this sample was sufficient to show specific and divergent effects of night shifts on two different cognitive constructs, i.e. sustained attention and RTB. Nevertheless, an enlargement of this sample would allow to consider a wider set of profiling variables, e.g. the chronotype,[67] as well as some confounding factors such as the presence of burn-out symptoms,[68] or the lack of specific training,[69] which have been associated to risky decisions. Finally, the lack of measures regarding working performance, patients’ safety and workplace injuries limits the translation of the current findings to real-life situations.

## CONCLUSIONS

Night shifts’ impact on risk-taking behavior and sustained attention in healthcare workers is modulated by different variables. In particular, sustained attention impairments follow sleep fragmentation during night shift, while risk-taking behavior increasing is specifically associated with individual characteristics, i.e. age and risk propensity. These results provide new insight on the link between night working and cognitive functioning in a medical context. Our findings suggest that the measures implemented to prevent performance decline during night shifts should consider the required cognitive processes, as well as vulnerability-specific workers’ profiling.

## Data Availability

The data underlying this article cannot be shared publicly for the privacy of individuals that participated in the study. The data will be shared on reasonable request to the corresponding author.

